# Computational Modeling of the Acoustic Radiation Force and Viscoelastic Displacement for Transcranial Focused Ultrasound Neuromodulation

**DOI:** 10.64898/2026.11.06.26367432

**Authors:** Tom Plovie, Emmeric Tanghe, Thomas Tarnaud

**Affiliations:** Department of Information Technology (INTEC-WAVES/IMEC), Ghent University/IMEC, Tech Lane 126, 9052 Zwijnaarde, Belgium; Department of Head and Skin (4Brain), Corneel Heymanslaan 10, 9000 Ghent, Belgium

## Abstract

**Objective:** In transcranial ultrasound neuromodulation (TUS), simulations of the harmonic pressure are often used for treatment planning. However, other mechanisms of energy transfer, such as acoustic radiation force (ARF) and the resulting tissue displacement, are reported less frequently. Furthermore, there is limited guidance on how these mechanisms should be simulated. This study evaluates the numerical convergence and accuracy of different ARF formulations and investigates the simulation of ultrasound-induced tissue displacement in the context of TUS.

**Approach:** Two simulations steps are used in k-Wave to first simulate the first order acoustic pressure and velocity fields. Subsequently, these fields are used to calculate the ARF using the momentum-flux density tensor, Reynold’s stress force, and attenuation force formulations. The ARF is then applied as input for a second viscoelastic simulation to estimate the tissue displacement induced by the ultrasound field. Numerical accuracy and convergence are quantified as functions of the various numerical choices and hyperparameters in the solvers.

**Main results:** Among the investigated formulations, the attenuation-force approach proved to be the most robust and the least susceptible to numerical errors across variable medium types and transducer geometries. Within the parameter space relevant to TUS, it is a valid approximation of the momentum flux density tensor formulation. Similar tissue displacement distributions are found across the different ARF formulations. However, the accuracy of the displacement simulation decreases with increasing pulse duration. Finally, recommendations are formulated on the ARF calculation method and on the numerical hyperparameters for ARF and displacement simulations.

**Significance:** This study offers guidelines for calculating the ARF and simulating tissue displacement. The proposed framework enables a more comprehensive characterization of ultrasound-mediated energy transfer in TUS.

## I. INTRODUCTION

Ultrasonic neuromodulation is a technique for non-invasive stimulation of neural tissues [1], [2]. Several physical mechanisms play a role in transferring energy from the transducer to the neurons [3]. Numerical simulations for treatment planning or computational studies for ultrasound neuromodulation generally concentrate on the harmonic pressure distribution [4], [5], [6]. However, acoustic radiation force (ARF) has been implicated as a main physical pathway for neuromodulation [7], [8], [9]. The ARF is a second order effect caused by the convective non-linearity in the Navier-Stokes equations, resulting in a non-oscillating force on the tissue [10]. In turn, ARF causes neural tissue displacement resulting in compression, elongation and bending of neurons. This causes perturbations of electrical properties like the capacitance and resistance of the neuronal membrane, as well as tension in the membrane that can cause activation of mechanosensitive ion channels [11], [12].

Regarding computational studies, the calculation methods of the harmonic pressure have been examined thoroughly. The influence of simulation parameters in the k-Wave tool-box like grid resolution are discussed in [13], showing the importance of fine grids to accurately model curved surfaces and to avoid the staircasing effect. Moreover, [14] investigated the sensitivity of the simulation result to medium parameters. A benchmark study demonstrated a maximal difference of 10 % in peak pressure and 1 mm in focal distance between 11 different numerical solvers for fine grid resolutions [15]. In [5], different solvers within k-Wave (`kspaceFirstOrder3D` and `pstdElastic3D`) are investigated, reporting a maximal difference of 15 %.

In contrast, there are only a limited number of numerical studies, investigating the ARF and displacement distribution for ultrasound neuromodulation. Various formulations of the acoustic radiation force have been proposed [10], [16], [17], [18]: momentum-flux density tensor force (MFDT), Reynold’s stress force (RS), and the attenuation force. [19] has shown the equivalency between the MFDT force and the attenuation force approximation for low attenuation co-efficients. In earlier studies, the ARF is primarily calculated by using a plane wave attenuation approximation [20], [21], [22]. These studies are mostly investigating elastography imaging. However, the accuracy of this approximation for neuromodulation is questioned in more recent studies [16], [17], [18]. Because of the lack of consensus, it is necessary to expand the research and to investigate how various formulations of the ARF perform in terms of numerical accuracy and computational requirements for typical transcranial ultrasound neuromodulation protocols.

Furthermore, the accuracy of ARF and displacement simulations is determined by various numerical hyperparameters: the voxel size (points per wavelength, PPW), the temporal step (set by the Courant-Friedrichs-Lewy number, CFL), the size of the perfectly matched layer (PML), and numerical choices for the calculation of the spatial gradients (k-space or finite difference derivatives). This study investigates through extensive parameter sweeps how the numerical settings and ARF formulation determine numerical convergence and accuracy of the radiation force and viscoelastic displacement, similar to [14] for pressure simulations. The goal of this study is to give guidance on the numerical settings for ARF and tissue displacement simulations for transcranial ultrasound neuromodulation.

## II. Methods

The general pipeline of the research is as follows. First, the pressure is simulated. From the pressure simulation, the ARF is calculated. Second, the ARF is used as input for the tissue displacement simulations. For both simulation steps, the influence of the most important simulation settings is investigated. The result of a simulation is quantified by multiple metrics, representing the characteristics of the respective field distributions. Ideally, the simulation settings reach a value for which further refinement no longer influences the field metrics (i.e., numerical convergence). For the calculation of the ARF, three formulations are tested and compared to each other as a function of the same simulation settings. Because of the prominent use of k-Wave for ultrasound neuromodulation simulations, this toolbox is chosen to investigate the aforementioned simulation features [23].

### A. Acoustic solvers and simulation software

All simulations are performed in MATLAB2022b, using the k-Wave toolbox [23]. k-Wave uses a pseudospectral solver, calculating the spatial and temporal derivatives in k-space and in time domain, respectively [23]. The equations that are solved are (conservation of momentum and mass):

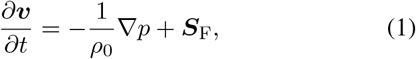

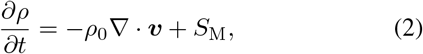

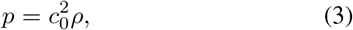

where ***v*** and *p* are the first order particle velocity and acoustic pressure, respectively. *ρ*_0_ is the equilibrium density, *c*_0_ is the speed of sound, *ρ* is the acoustic density, ***S***_F_ is the force source term, and *S*_M_ is the mass source term.

For the simulation of tissue displacement, the elastic solver of k-Wave is used (`pstdElastic3D` solver) [24]. This solver accounts for the propagation of compressional and shear waves in isotropic viscoelastic solids. The equations that are solved in this case are (conservation of momentum and Kelvin-Voigt model; Einstein summation notation is used):

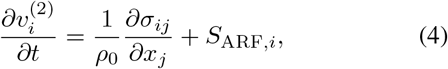

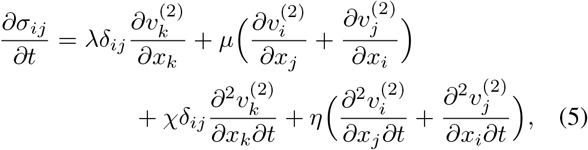

where ***v***^(2)^ is the second order particle velocity, *S*_ARF,*i*_ the ARF source term (cf. section II-E), ***σ*** is the Cauchy stress tensor, and *δ*_*ij*_ is the Kronecker delta function. *λ* and *µ* are the Lamé parameters:

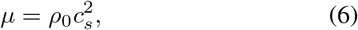

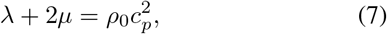

where *c*_*s*_ is the shear velocity, and *c*_*p*_ = *c*_0_ is the compressional velocity. Furthermore, *χ* and *η* are the compressional and shear viscosity coefficients, given by

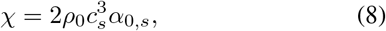

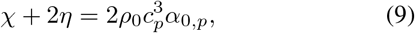

where *α*_0,*s*_ is the shear wave attenuation and *α*_0,*p*_ is the compressional wave attenuation [24], with 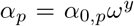 (*y* is a power law exponent, *ω* is the angular frequency).

## B. Pressure simulations

The pressure simulations are performed in an axisymmetric (`kspaceFirstOrderAS` solver) and in a 3D setup (`kspaceFirstOrder3D` solver) [25], [23]. For both setups, different simulation domains are used. The simulation domains represent ultrasound propagation in free space and through a half-spherical homogeneous skull for the axisymmetric setup. For the 3D setup, one additional simulation domain is used, which includes a skull CT scan. The CT scan is selected from the cq500 dataset (model ID: 100) [26]. The domains are shown in Figure 1. The sizes of the domains are 40 mm × 40 mm × 52 mm, 36 mm × 36 mm × 74 mm, and 36 mm × 36 mm × 111 mm for free space, half spherical skull, and realistic skull, respectively.

**Fig. 1:**
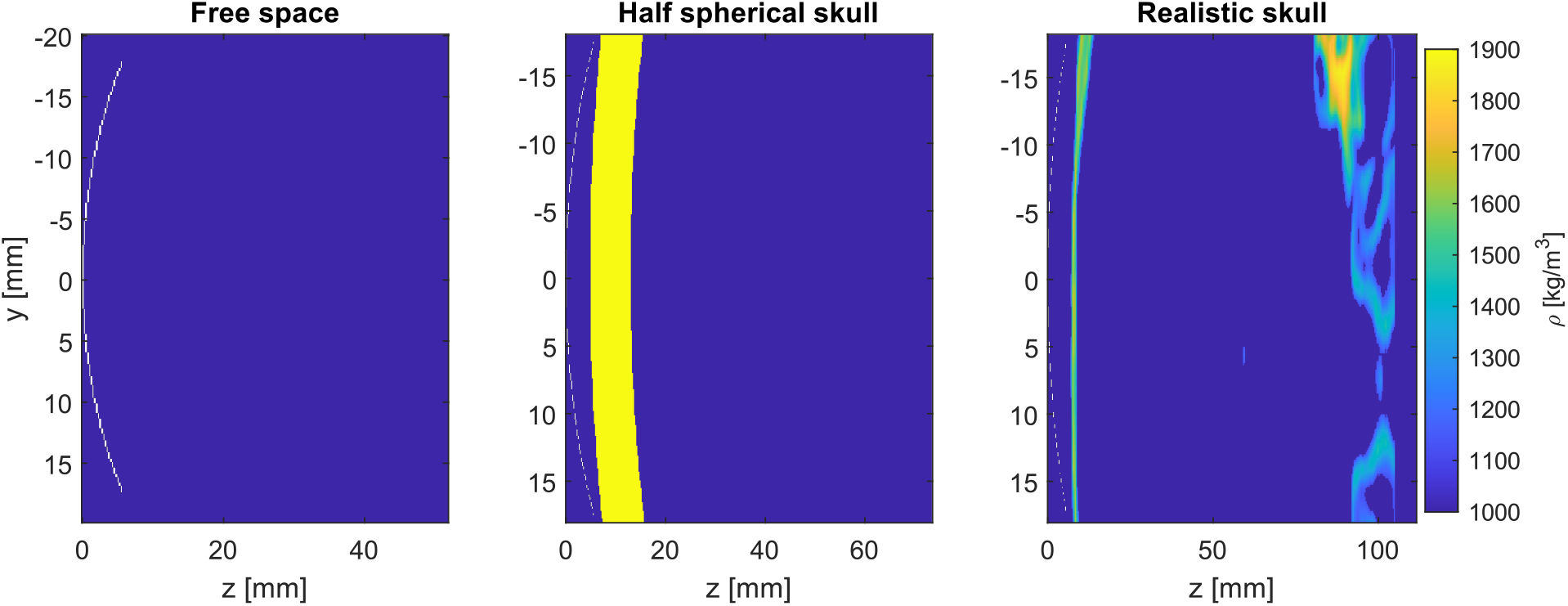
Simulation domains used in this study (left to right): free space, half-spherical skull, realistic skull. The transducer is placed at the left side (white) of the domain pointing to the right. A perfectly matched layer (PML) is added around the whole domain (not shown).

The medium properties used for the free space and the half spherical skull simulation are given in Table I. Note that there are no shear wave parameters for the skull, since the skull is left out in the displacement simulation to reduce the computation time (cf. section II-E). The thickness of the half-spherical skull is set to 8 mm to represent the human skull [27], [28], [29]. The radius of curvature of this skull is set to 70 mm [30]. For the CT skull, the medium properties are calculated from the Hounsfield units (HU). The HU describe the radiodensity of the CT scan and are defined as:

**Table I:** Medium parameters.

| **Quantity** | **Value** | **Reference** |
|---|---|---|
| α<sub>p,brain</sub> | 3.6 Np/m | [17] |
| α<sub>s,brain</sub> | 3.6 Np/m | — |
| ρ<sub>0,brain</sub> | 1000 kg/m<sup>3</sup> | [17], [19] |
| c<sub>p,brain</sub> | 1500 m/s | [17], [19] |
| c<sub>s,brain</sub> | 2 m/s | [36] |
| α<sub>p,skull</sub> | 25 Np/m | [31] |
| ρ<sub>0,skull</sub> | 1900 kg/m<sup>3</sup> | [31] |
| c<sub>p,skull</sub> | 2800 m/s | [31] |

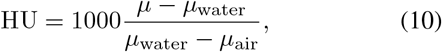

where *µ* is the x-ray attenuation parameter. The HU are calculated from the pixel unit by first adding the slope of the dicom image, which is 1024 in this case. The conversion formulas are:

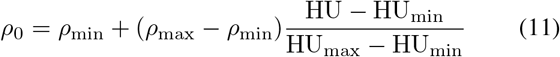

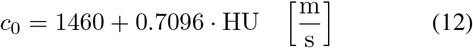

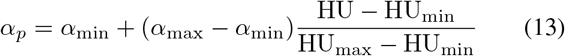

with *ρ*_min_ = 1000 kg*/*m^3^, *ρ*_max_ = 1900 kg*/*m^3^, 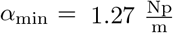, and 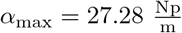 [31]. Multiple models have been proposed to translate HU to medium properties. Here, we use conversion formulas from [32], [33], [34].

### C. ARF formulation

Three formulas of the acoustic radiation force are examined ((14)-(16)). Because both axisymmetric and 3D simulations are performed (i.e., cylindrical and Cartesian coordinates), the numerical implementation of the formulations is adjusted accordingly. The derivation of the ARF formulations is given in [18]. The momentum flux density tensor (MFDT) formulation is given by:

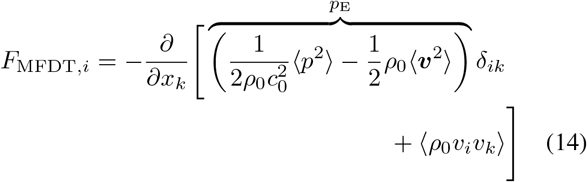

Here, ⟨·⟩ refers to the time average over the acoustic period. When the first term (Eulerian excess pressure *p*_E_) is dropped out, this is called the Reynold’s stress (RS) formulation as shown in (15). The Eulerian excess pressure is a gradient term in the driving force of the viscoelastic displacement equations (4)-(5). As a result, although the Reynold’s stress and MFDT ARF distributions will be different, both formulations are expected mathematically to result in identical viscoelastic displacement distributions under the assumption of incompressibility, except for numerical errors (cf. supplementary information).

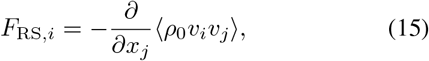

It is shown in [35] for low attenuation, that the MFDT formulation can be simplified to:

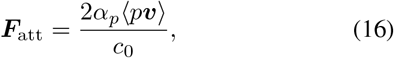

where *α*_*p*_ is the attenuation. This is the attenuation formulation. Here, we note that the vectorial nature of the attenuation force in (16) is maintained and that no further plane wave simplification is applied (*p* = *ρ*_0_*c*_0_*v*). I.e., the pressure *p* and velocity ***v*** are calculated independently with the `kspaceFirstOrder`-solvers (eqs. (1)-(2)).

### D. ARF calculation method

Two different aspects are investigated with respect to the calculation method of the MFDT and RS formulations of the ARF. First of all, the derivatives are calculated with the following two methods (gradient and k-space derivative):

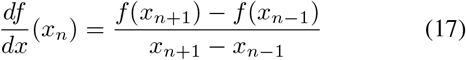

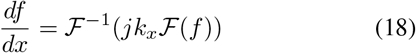

Here, ℱ and *k*_*x*_ refer to the Fourier transform and the x-component of the wavenumber, respectively.

Second, the influence of reversing the order of calculating the mean and taking the derivative is examined because both can be found in literature [19], [17]. The difference is shown below. The equations for examining the order of operations are shown in cylindrical coordinates because the simulations for this part are performed using the axisymmetrical solver. For the Reynold’s force, calculating the mean first (m1):

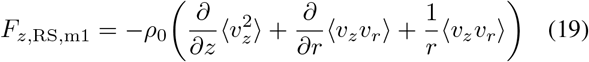

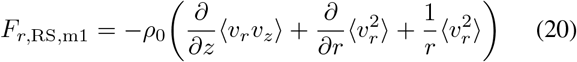

The formulas of the Reynold’s force, calculating the derivative first (d1) are:

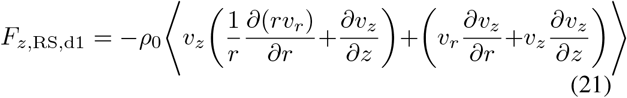

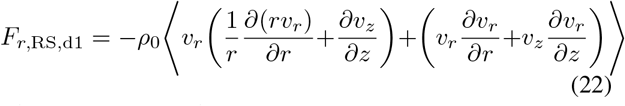

The equations for the MFDT formulation are analogous.

### E. Displacement simulations

The displacement of brain tissue is simulated by solving (4)-(5) in k-Wave, with the viscoelastic properties in Table I. The compressional speed of sound and attenuation are the same as in the pressure simulations. The shear wave speed in the brain is 2 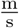 [36]. The shear wave attenuation of the brain is set equal to the compressional wave attenuation because its precise value does not impact the final result. This is shown in the supplementary information. The skull is not included in the displacement simulations. This is also further explained in the supplementary information. Simulations are performed for both a continuous input of 80 ms (free space), as well as a pulsed wave of 5 ms, where the simulation is ran for 10 ms (realistic skull).

The ARF is used as an input for the displacement simulation. In k-Wave, the force source is defined as

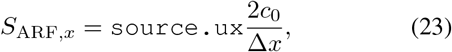

where `source.ux` is the source velocity in the x-direction and Δ*x* is the voxel size.

In order to do so, the input velocity is calculated as 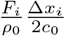 . This is inputted in the `pstdElastic3D` solver of k-Wave [24]. Since the grids are not the same for the pressure simulation and the displacement simulation, the ARF is linearly interpolated to be used as an input. The voxel size of the displacement simulation is kept larger than the voxel size of the pressure simulation in order to minimize numerical errors from interpolation. From this simulation, the second order particle velocity is retrieved. Similar as in [35], this particle velocity is integrated over time to determine the tissue displacement. The domain sizes for the displacement simulations are 3 mm × 3 mm × 25 mm and 7 mm × 6 mm × 25 mm for free space and realistic skull, respectively.

### F. Default parameters and sensitivity analysis

The parameter values for the baseline pressure simulation are summarized in Table 2. The ultrasound frequency is set to 500 kHz. The amplitude of the surface pressure of the transducer is set to 1 MPa. The transducer is placed at (*y, z*) = (0, 0) mm. Because of the linearity of the solvers, the results are valid for all pressure amplitudes: first order particle velocity and pressure scale linearly with transducer pressure, while second order velocity, tissue displacement, and ARF are quadratic with the transducer pressure. Regarding the transducer properties, two sweeps are performed to examine the influence of radius of curvature and aperture on the numerical convergence. The attenuation coefficient is investigated over a range from 1 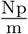 to 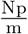 .

**TABLE II:** Default values and parameter ranges of the simulation setup for pressure and ARF calculation, except when explicitly mentioned otherwise. Heterogeneous denotes all simulations including skull tissue.

| Quantity | Value | Tested range |
|---|---|---|
| CFL (homogeneous) | 0.3 | 0.05-0.3 |
| CFL (heterogeneous) | 0.05 | 0.01-0.05 |
| PML (homogeneous) | 5 mm | 0.2 mm - 10 mm |
| PML (heterogeneous) | 10 mm | 0.2 mm - 40 mm |
| PPW (homogeneous / axisymmetric) | 12 | 5-40 |
| PPW (homogeneous/3D) | 12 | 5-25 |
| PPW (heterogeneous) | 17 | 5-25 |
| Radius of curvature | 30 mm | 30 mm - 60 mm |
| Aperture | 35 mm | 31 mm - 60 mm |
| Order of operations | Mean first | Mean vs derivative first |
| Derivative | Gradient | Gradient vs k-space |

For the displacement simulations, a sensitivity analysis is performed for both the spatial grid and the time step. Similar grid parameters to the pressure simulations are used like ppw, PML size, and CFL. An additional parameter is the buffer size. This is a zone that is added between the PML and the source space (= ARF field) to assure that the sources are further from the edges of the simulation domain. The reason to add this extra parameter is because it became clear in the displacement simulations that inclusion of a buffer can reduce the accumulation of numerical noise for long pulse durations. The investigated parameter ranges are summarised in Table 3. To limit the computation time in the displacement simulations, the default parameters are not the same across the sweeps of different parameters.

**TABLE III:** Parameter settings for displacement simulation sweeps.

| Variable parameter | Simulation case | PPW [-] | CFL [-] | PML [mm] | Buffer [mm] |
|---|---|---|---|---|---|
| PPW | Free space, continuous | 5-10 | 0.3 | 3 | 3 |
|  | Real skull, 5 ms pulse | 5-8 | 0.3 | 5 | 7 |
| CFL | Free space, continuous | 6 | 0.2-0.3 | 5 | 10 |
|  | Real skull, 5 ms pulse | 6 | 0.15-0.3 | 5 | 7 |
| PML | Free space, continuous | 6 | 0.3 | 3-11 | 10 |
|  | Real skull, 5 ms pulse | 6 | 0.3 | 3-11 | 7 |
| Buffer | Free space, continuous | 6 | 0.3 | 5 | 3-13 |
|  | Real skull, 5 ms pulse | 6 | 0.3 | 5 | 1-9 |

### G. Field distribution metrics

The results are analysed both visually and by looking at more specific metrics. For all simulations, the maximal force, the distance from the ARF focus to the transducer, and the full width at half maximum (FWHM) in the axial direction (i.e., the z-direction) are calculated based on the ™ 3 dB points of the ARF inside the brain tissue. Here, the ARF focus is defined as the location in the brain corresponding to the maximum ARF. For the heterogeneous simulations, the maximum inside the brain tissue is taken, although a higher ARF might be present inside the skull. For the convergence analysis, the result with the highest grid resolution is taken as the ground truth, to which the lower-resolution simulations are compared. Moreover, the regions where the ARF increases towards the border of the simulation domain are left out of the calculation of the FWHM because they are seen as an artifact of the simulation due to periodic boundary conditions.

For the displacement simulation, similar metrics are extracted: maximal displacement, focal distance, and FWHM of the displacement distribution. Additionally, the rise time is calculated. This is defined as the time for the spatial peak displacement to increase from 10% to 90% of the maximum. Depending on the context, the terms focal distance and FWHM refer to these metrics calculated for the pressure, ARF or displacement distribution.

## III. Results

### A. Courant number does not impact simulation accuracy of stable simulations

The Courant-Friedrichs-Lewy number (CFL) is a dimensionless number, determining the time step and is given by

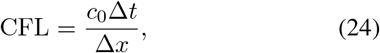

where, Δ*t* is the time step, and Δ*x* is the voxel size. The effect of the CFL number is minimal. For free space, the simulations are tested for a CFL from 0.05 to 0.3. For the realistic skull, the CFL is tested from 0.01 to 0.05 (TABLE II). All simulations were performed with the 3D solver. For all CFL values and ARF formulations (attenuation, MFDT, RS), the maximal force, the focal point and the FWHM are analysed. The definitions of these metrics are shown in Figure 2, together with an example of the pressure, ARF and displacement distribution in a plane (Figure 2A-C) and on the main axis (Figure 2D-F). In free space, the difference of the maximal force between the largest and smallest CFL is less than 1 % for all formulations. For the focal distance and FWHM there is no difference within the resolution of the grid (= 0.25 mm). For the realistic skull, the difference in maximal force is 1.1 % between a CFL of 0.01 and 0.05 for the attenuation force. The difference is less than 1 % for the other formulations. For the other metrics, the only notable difference is a 2.2 % difference of the FWHM in the attenuation force between a CFL of 0.01 and 0.05. It should be noted that when the CFL becomes larger than 0.3 in free space and larger than 0.05 for the realistic skull, the simulation quickly becomes unstable. Thus, it can be concluded that as long as the simulation is stable, the exact choice of the CFL does not matter. As the simulation time rises with lower CFL, the CFL is chosen as 0.3 for the homogeneous case and 0.05 for heterogeneous cases.

**Fig. 2:**
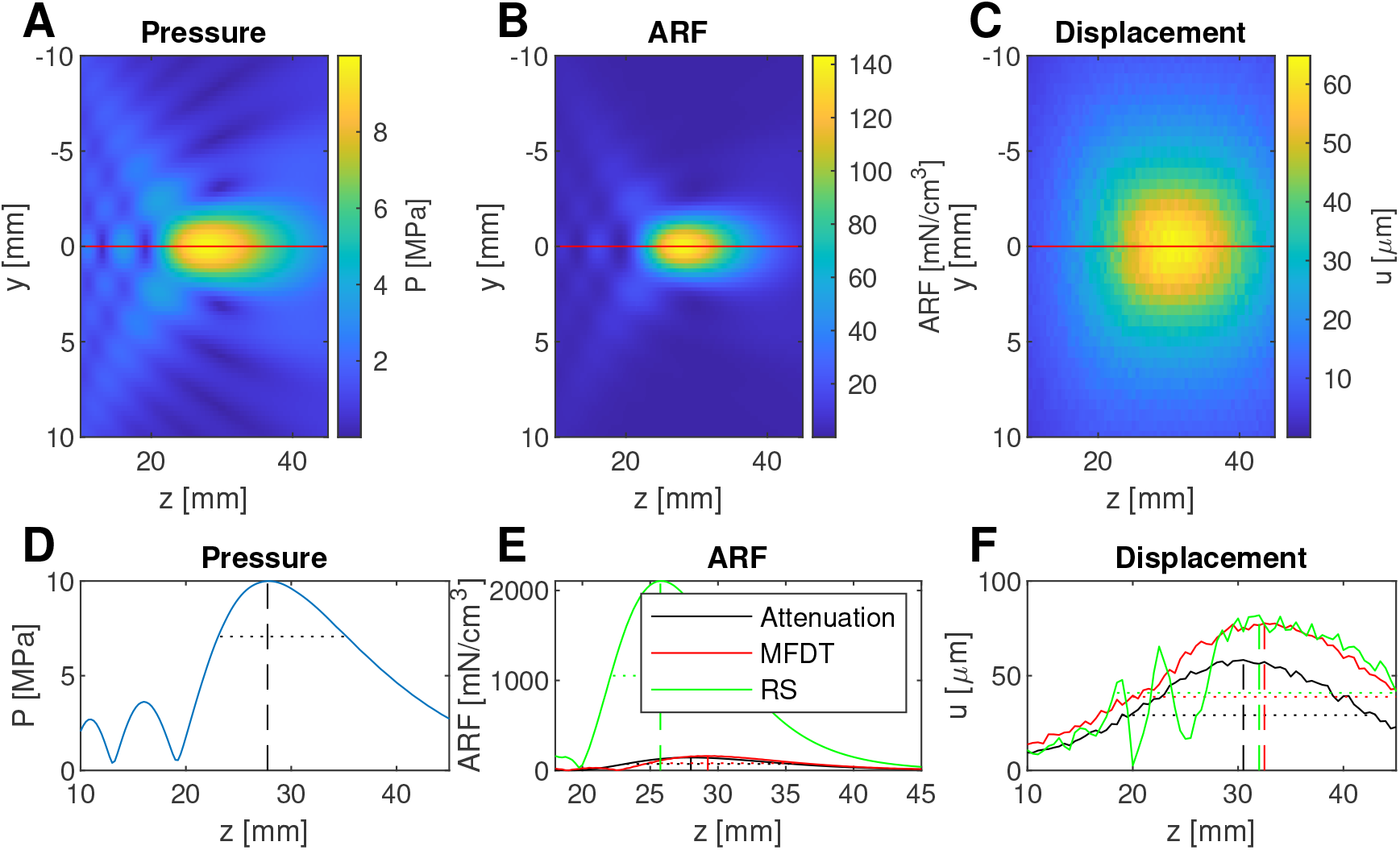
Illustration of the metrics of the field distributions. **A-C**: 2D colour maps of pressure, ARF magnitude (attenuation formulation), and tissue displacement. **D-F**: 1D profiles through the maximum (red line in **A-C** for the attenuation formulation) showing the focal distance (vertical dashed line) and the FWHM (dotted horizontal line). Pressure simulations are performed in free space with the 3D solver.

### B. Minimal thickness of the perfectly matched layer

The perfectly matched layer (PML) is added at the boundary of the simulation domain to mimic an infinite space. In the case of k-Wave, it prevents the acoustic waves from entering on the other side of the simulation domain when reaching the edge of the domain due to the periodic boundary conditions. The effect of the PML is shown in Figure 3. These simulations were also performed with the 3D solver. It is clear that the PML needs a minimal thickness in order to minimize waves propagating through the simulation domain borders. In free space, the changes are smaller than 2 % from a PML of 0.6 mm on for all formulations and for all metrics (Figure 3A-C).

**Fig. 3:**
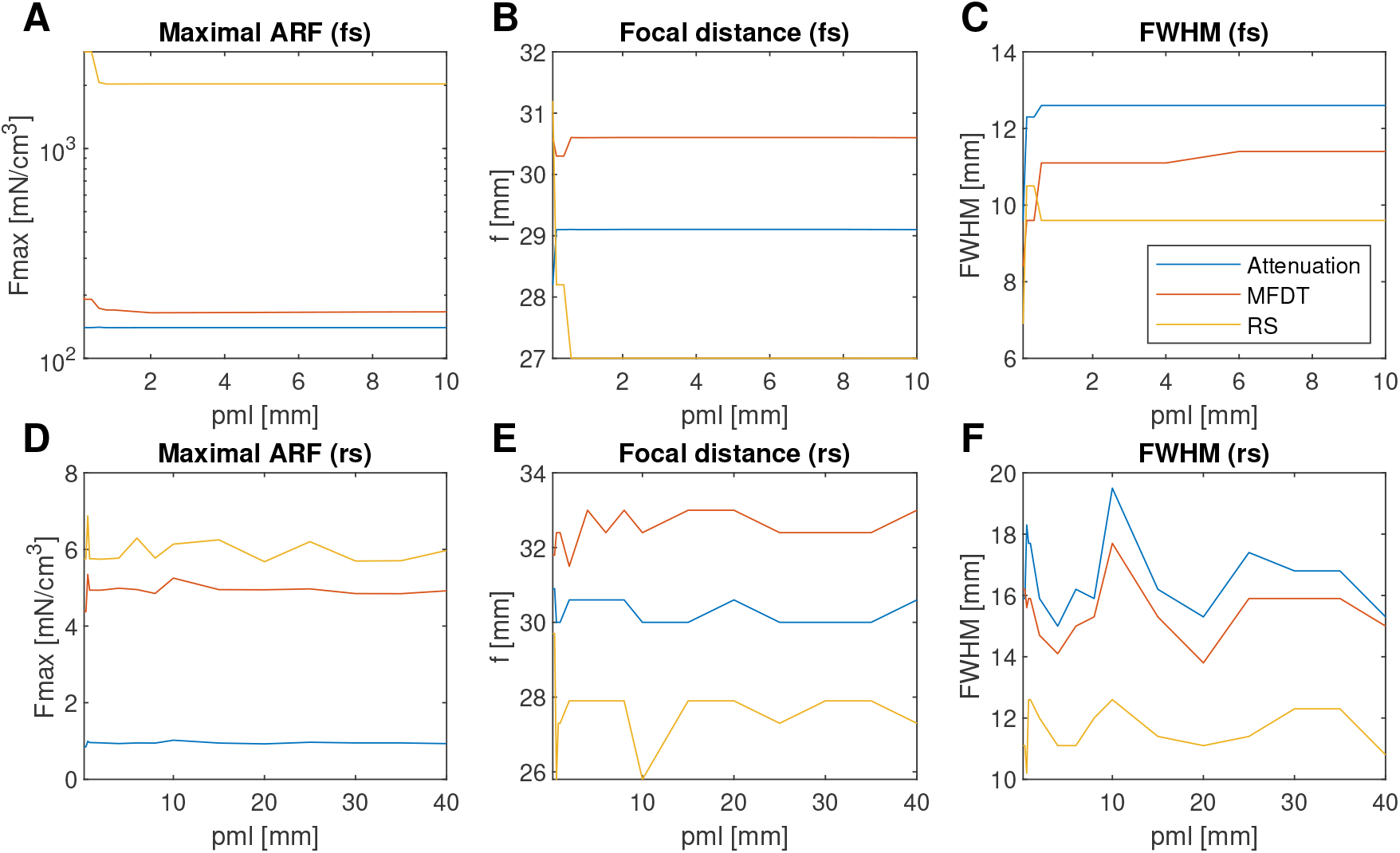
Influence of the PML showing convergence from 2 mm on for the homogeneous case (fs: free space) for all formulations and from 10 mm on for the heterogeneous case (rs: realistic skull). (**A**) Maximal ARF magnitude, (**B**) focal distance, and (**C**) FWHM for the homogeneous simulation. (**D**) Maximal ARF magnitude, (**E**) focal distance, and (**F**) FWHM for the heterogeneous simulation. All metrics are calculated on the main axis of the transducer.

For the realistic skull, there is more variation with changing PML thickness (Figure 3D-F). For the attenuation force, the biggest difference in amplitude from the maximal PML size of 40 mm is 9.5 % for PML sizes greater than 1 mm. However, this seems to be caused by an outlier at PML = 10 mm, for which the spatial oscillations are more prominent, as all other maximal ARF values have a difference of 3.8 % or less.

From 0.6 mm on, the maximal ARF and focal distance of all formulations are within an error of 10 %. For the FWHM, the impact of the PML size is more pronounced with some cases where the error is higher than 10 % (e.g., all formulations at a PML size of 10 mm and attenuation force at a PML size of 25 mm).

From the results in Figure 3, it can be concluded that for the homogeneous case, a PML size of at least 2 mm is necessary to get all metrics below 2 % difference withrespect to the simulation with the thickest PML. In further simulations, a PML of 5 mm is chosen. On the other hand, for the heterogeneous case, no real convergence is seen as in the homogeneous case, but for a PML of at least 4 mm, the numerical error on all metrics is below 10 %, apart from a few exceptions. In further heterogeneous simulations, a PML size of 10 mm is chosen.

### C. First calculating mean and afterwards the derivative assures faster convergence

The results of the influence of the order of operations is shown in Figure 4. Here, an axisymmetric solver is used to simulate higher grid resolutions than would be computationally feasible with the 3D solver. Because of the singularity on the main axis for the axisymmetric case, the result is plotted and analyzed for *r* = 1 mm. It can be seen that both methods eventually give similar results for high grid resolutions. However, there is a big difference for smaller grid resolutions. In free space for 5 ppw, the method of first calculating the derivative gives an overestimation of 800 % for the MFDT formulation and an underestimation of 9.6 % for the RS formulation in terms of the ARF magnitude. When first calculating the mean, the over- and underestimation is 140 % and 25 %, respectively. The trend for the MFDT formulation is observed in general: over the range of tested grid resolutions, taking the mean first results in a lower error than taking the derivative first for all metrics and both for the free space and half-spherical skull domains. The focal distance and FWHM do not fully converge when taking the derivative first. Additionally, in Figure 4C for the heterogeneous domain, strong oscillations can be seen when taking the derivative first. For the RS formulation it is the other way around: the error is larger when taking the mean first. Here, in contrast to the MFDT formulation, both calculation methods do converge.

**Fig. 4:**
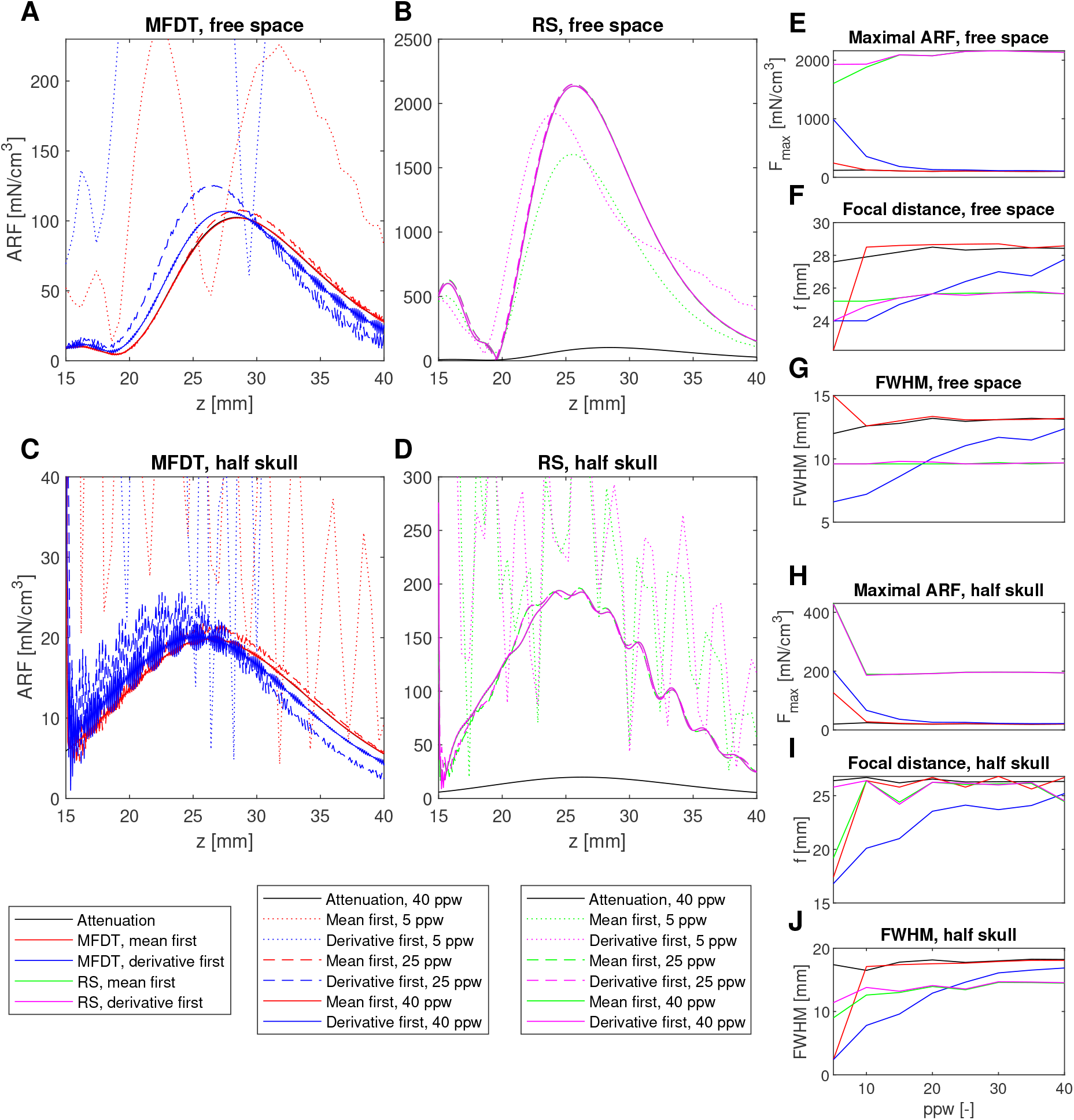
Influence of order of operations on convergence of ARF calculation for axisymmetric simulations. **A-D**: ARF magnitude. **E-G**: ARF magnitude, focal distance, and FWHM for homogeneous case. **H-J**: ARF magnitude, focal distance, and FWHM for the heterogeneous spherical skull case. All metrics are calculated on an axis parallel to the main axis of the transducer with an offset of 1 mm.

To conclude, for the MFDT formulation it is beneficial to take the mean first. For the RS formulation it could be advantageous to calculate the derivative first, although in general it still seems acceptable to calculate the mean first.

### D. K-space derivative assures faster convergence, but with stronger oscillations

The second aspect of the calculation method of the MFDT and RS formulations is the numerical method that is used to calculate the derivative. Two methods are compared that are described in section II-D: calculating the gradient versus taking the derivative in k-space ((17) vs (18)). This aspect of the calculation is investigated for a full 3D simulation to avoid misinterpretations due to the singularity at *r* = 0 in the derivative using the gradient method, which is not present in the k-space derivative. The results are shown in Figure 5. For free space, it is clear that the k-space method converges much faster than the gradient method. For the k-space method, already from 5 ppw on, the result looks comparable with the result of 25 ppw. At 7 ppw, the numerical error on the maximal ARF, focal distance and FWHM is already below 5 % for both formulations. For the calculation using the gradient it takes until 12 ppw to have a numerical error lower than 10 % for all metrics. At 25 ppw, the difference in maximal ARF between both calculation methods is 4.1 % for the MFDT formulation and 0.8 % for the RS formulation. One issue with the k-space method is that oscillations stay present for all grid resolutions. These are mostly observed for the MFDT formulation and are most prominent closer to the domain edges. This is a result of the Gibbs phenomenon, causing oscillations that become more prominent when calculating the derivative in k-space [37], [38].

**Fig. 5:**
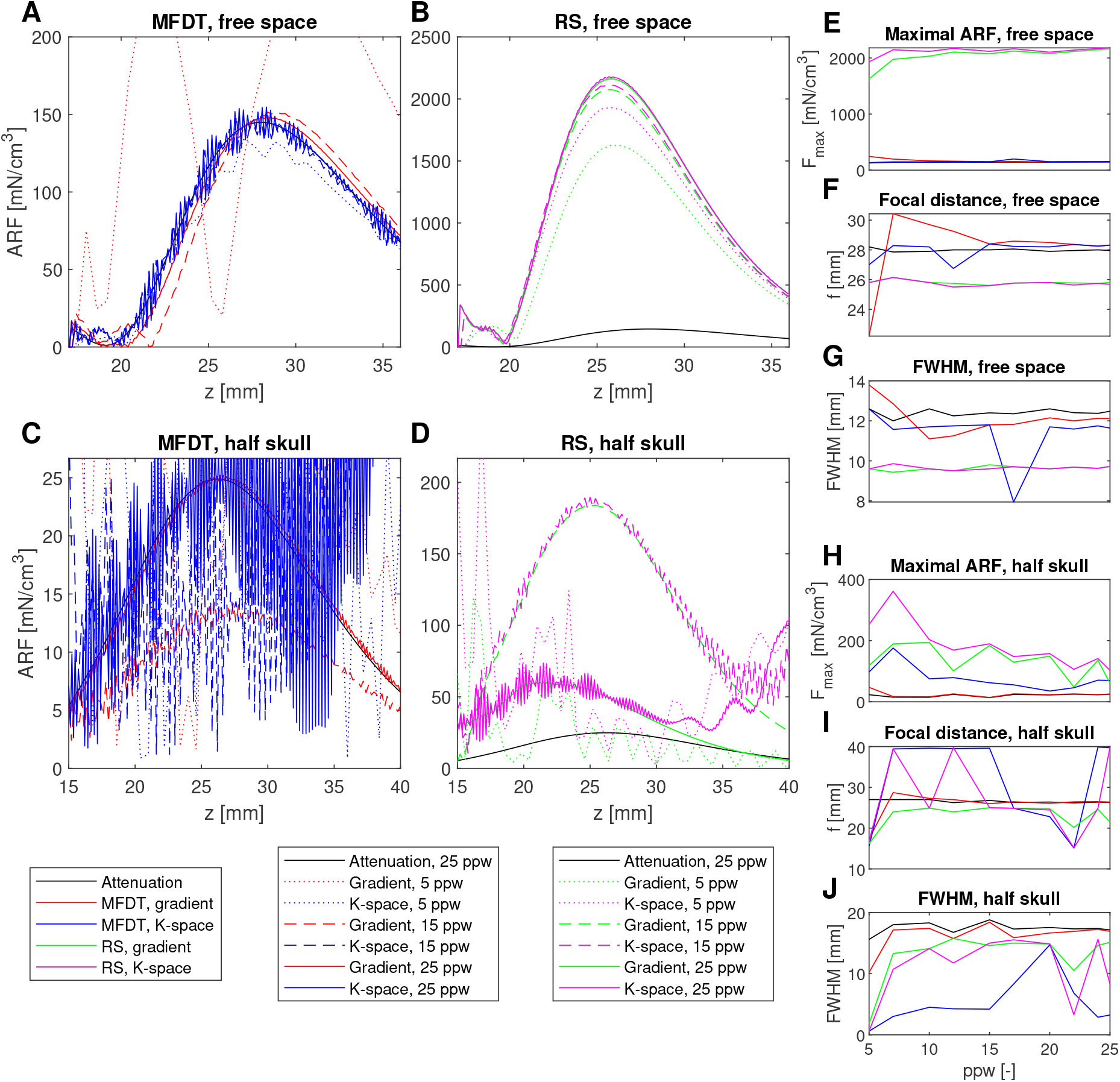
Influence of the derivative method on convergence of ARF calculation for 3D simulations. **A-D**: ARF magnitude. **E-G**: ARF magnitude, focal distance, and FWHM for the homogeneous case. **H-J**: Total ARF, focal distance, and FWHM for the heterogeneous spherical skull case. All metrics are calculated on the main axis of the transducer.

For the half-spherical skull, the problem of the Gibbs phenomenon becomes more pronounced. Especially the convergence plots of the focal distance and the FWHM show big jumps because of the oscillations (Fig. 5I-J). Without additional techniques to reduce Gibbs artifacts, it seems not possible to apply the k-space derivative to heterogenous media for the calculation of the acoustic radiation force, especially for the MFDT formulation. For the gradient method in the MFDT formulation, at least 17 ppw is necessary to keep all metrics below a difference of 10 % compared to 25 ppw.

To keep consistency and to avoid the oscillations in free space as well, the gradient method is used for further simulations. Note that there is a difference between the ARF in Figure 4 and 5. This is due to the offset (1 mm) with respect to the x-axis in the axisymmetric case due to the singularity at the main axis of the transducer.

### E. Attenuation formulation converges at coarser grids than MFDT and RS formulation

A comparison of the results for all three ARF formulations as a function of grid resolution, for different media and a single transducer geometry, is shown in Figures 4 and 5. The distributions in Figures (4-5)A&C, as well as the quantitative metrics in Figures (4-5)E–J, show that the MFDT and attenuation formulations produce very similar results. The quantitative results at a grid resolution of 25 ppw are summarized in Table IV. For both the free-space and half-skull simulations, the MFDT formulation and the attenuation formulation match closely in terms of both maximum force and focal distance. In contrast, the Reynolds stress (RS) formulation predicts substantially larger maximum forces and a noticeably shorter focal distance.

**Table IV:**
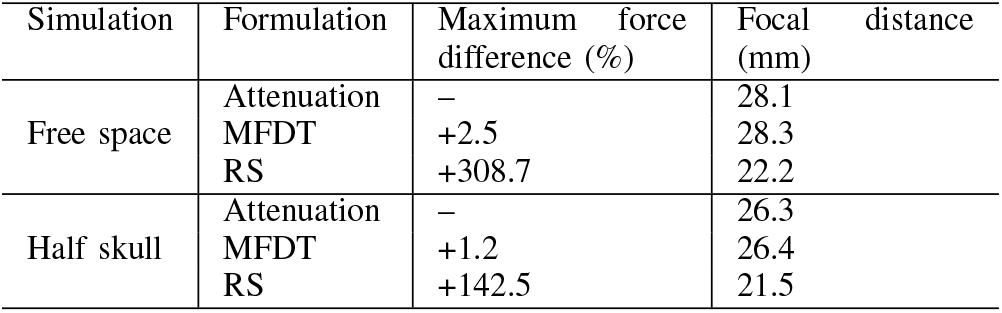
Comparison of ARF formulations at 25 ppw for the 3D solver. Percentage differences in maximum force are reported relative to the attenuation formulation.

Furthermore, the attenuation and MFDT formulation converge at the same speed for the half-spherical skull: both need 17 ppw to have all metrics below 10 % error (axisymmetric and 3D solver). In free space, the attenuation formulation converges faster: only 7 ppw are needed to get below 10 % error while the MFDT formulation needs 12 ppw. However, the attenuation and MFDT distributions differ significantly for the realistic skull simulations, for which reflections at the back of the skull are possible (Figure 6G-H). In this case, the pattern of the MFDT qualitatively looks more like the RS formulation (Figure 6I). When the simulation domain becomes more complex, there is thus again more difference between the attenuation formulation and the MFDT formulation. The reason might be that the first term of the MFDT formulation ((14), Eulerian excess pressure) converges slower for more complex media, hence the better correspondence with the RS formulation.

**Fig. 6:**
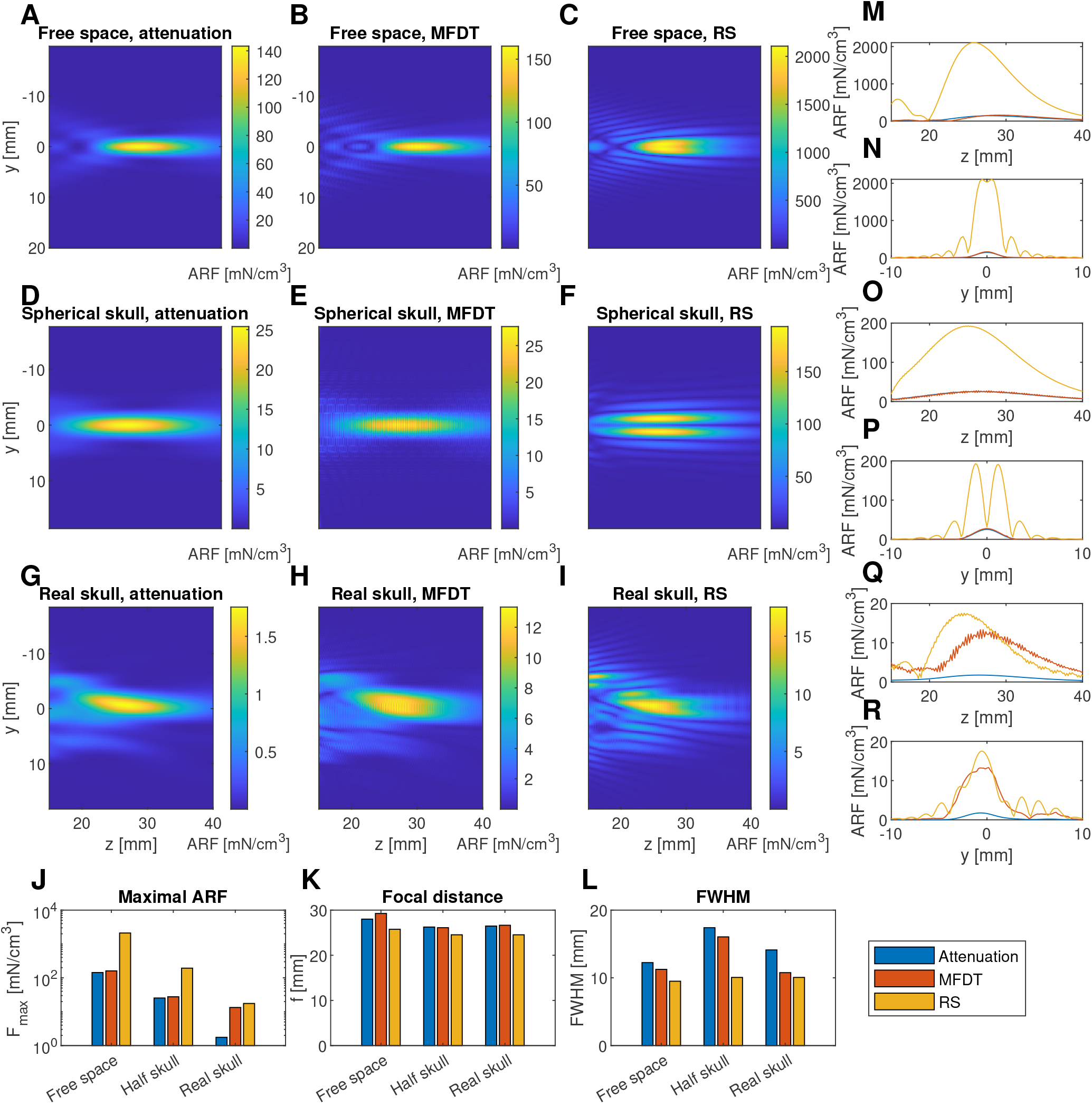
Effect of the formulation of the ARF for 3D simulations. **A-C**: ARF in homogeneous medium for attenuation, MFDT, and RS formulation. **D-F**: ARF in heterogeneous medium with half spherical skull for attenuation, MFDT, and RS formulation. **G-I**: ARF in heterogeneous medium with realistic skull for attenuation, MFDT, and RS formulation. **M-N**: one-dimensional plots of the ARF in homogeneous medium in axial direction and lateral direction, respectively. **O-P**: one-dimensional plots of the ARF in heterogeneous medium with half spherical skull. **Q-R**: one-dimensional plots of the ARF in heterogeneous medium with realistic skull. **J-L**: bar plots showing the maximal ARF, focal distance, and FWHM for all ARF formulations in all media. All metrics and one-dimensional plots are for an axial or lateral axis through the maximum. The legend used in J-L and M-R is shown in the bottom-right.

For the real skull simulations at 17 ppw, the maximal ARF differences of the MFDT formulation and the RS formulation are 660 % and 899 %, respectively, w.r.t the attenuation formulation. The focal distances are 26.5 mm, 26.6 mm, and 24.5 mm for the attenuation, MFDT, and RS formulation, respectively. Again, some oscillations can be seen for the MFDT formulation in Figure 6. They are kept to a minimum because of the choice to take the derivatives through the gradient instead of the k-space derivative.

Thus, the attenuation formulation converges fastest which can be explained by the fact that no derivatives are needed. Moreover, the attenuation formulation is much less susceptible to the Gibbs phenomenon because of the lack of spatial derivatives, which especially affects the heterogeneous simulations. Because of this, it is recommended to choose the attenuation formulation for the calculation of the ARF.

### F. The attenuation only affects the simulation precision at low values

The effect of the attenuation coefficient on the convergence of the ARF is investigated for the case of a homogeneous medium in Figure 7. The attenuation force and the MFDT force are more similar for higher attenuation coefficients. For *α*_*p*_ = 1 Np*/*m, the difference between the attenuation force and the MFDT force is less than 5% from 20 ppw on. For *α*_*p*_ = 2 Np*/*m, the difference between attenuation force and MFDT force is lower than 5% for all grid resolutions except for 11 ppw. For *α*_*p*_ = 5 Np*/*m and *α*_*p*_ = 10 Np*/*m, the difference is always less than 5%.

**Fig. 7:**
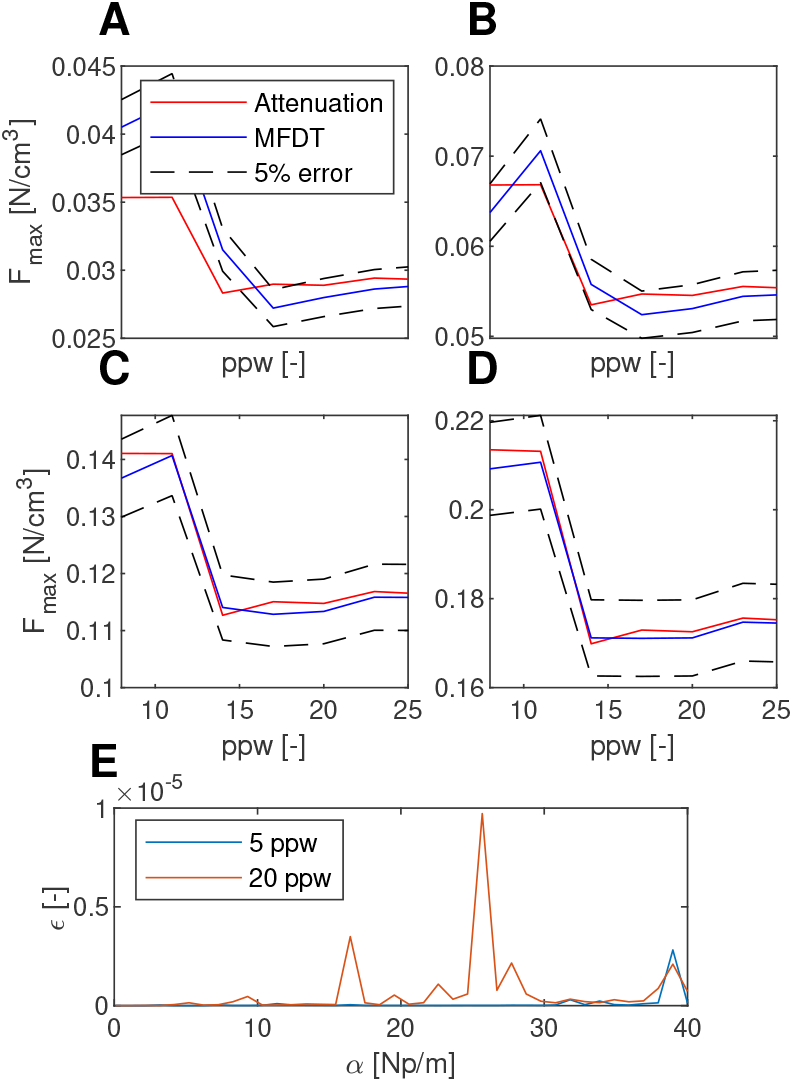
Effect of the attenuation on the ARF convergence in the axisymmetric solver (homogeneous medium, maxima on axis with offset of 1 mm with respect to the main axis). **A-D**: Maximal ARF for *α*_*p*_ = 1 Np*/*m, *α*_*p*_ = 2 Np*/*m, *α*_*p*_ = 5 Np*/*m, and *α*_*p*_ = 10 Np*/*m, respectively. **E**: Ratio of second order to first order attenuation (*ϵ*) showing the second order term of the attenuation is negligible for all cases.

This observation seems to contradict [19] which states that the attenuation formulation and the MFDT formulation are mathematically equivalent for low attenuation coefficients. However, numerical errors increase for lower attenuation coefficients, explaining the larger discrepancies between both formulations in Figure 7A-B. According to [19], the attenuation is low when the second order attenuation term is small enough (derivation shown in the supplementary information):

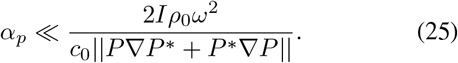

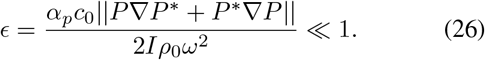

*I* is the ultrasonic intensity. Here, phasor notation is used for the pressure *P* and its complex conjugate *P* ^*^ (with sign convention: *p* = ℜ (*Pe*^™*jωt*^)).

This comparison is checked for a grid resolution of 5 ppw and 20 ppw, for 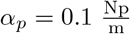to 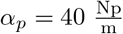 . The condition in (26) is satisfied over the whole range (Figure 7E).

### G. Radius of curvature has bigger influence on convergence than aperture

The influence of the transducer geometry on the convergence of the ARF is shown in Figure 8. The reference case is a transducer with a radius of curvature (roc) and an aperture (ap) of 60 mm. It can be seen that in all cases except for the radius of curvature of 31 mm, the attenuation force and the MFDT force are within a margin of error of 5 % of each other between 8 ppw and 26 ppw.

**Fig. 8:**
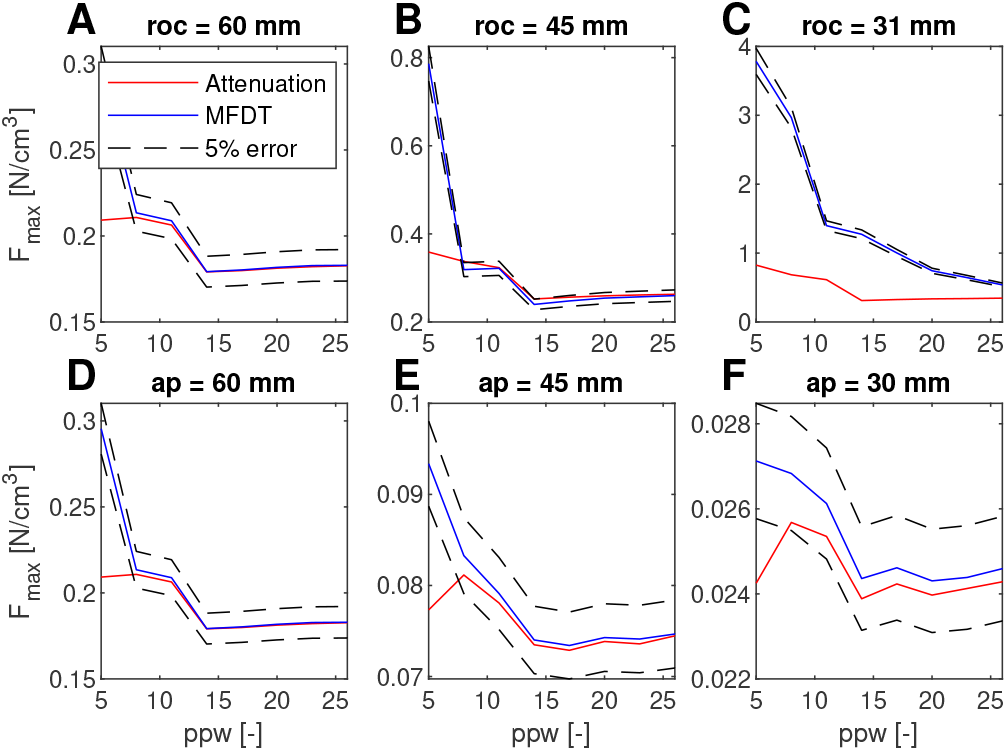
Effect of the transducer geometry on the ARF convergence in the axisymmetric solver (homogeneous medium, maxima on axis with offset of 1 mm with respect to the main axis), especially showing the influence of the radius of curvature on the convergence of MFDT and attenuation force towards each other. In **A**-**C**, the aperture is kept constant at 60 mm. In **D**-**F**, the radius of curvature is kept constant at 60 mm.

The impact of the transducer aperture on the ARF convergence is less prominent. For all cases, the relative error between the MFDT and attenuation force between 8 ppw and 26 ppw is below 2 % and decreases with larger apertures.

Both formulations converge equally fast with respect to the grid resolution for all transducer geometries except for the roc of 31 mm. In this case, the attenuation force has an error below 10 % w.r.t. the most precise attenuation force simulation from 14 ppw on, while the MFDT force does not fully converge. This indicates that for this case, the attenuation force formulation is a better way to calculate the ARF than the MFDT formulation at low grid resolutions.

### H. Displacement simulations

#### 1) Continuous, free space

The displacement simulation is performed for an ARF input related to a maximal pressure of 10 MPa (Figure 2D). Displacement profiles for other input pressures can be obtained directly from these results by quadratic scaling with input pressure. The investigated parameter ranges are given in TABLE III. Note that because the default parameters are not the same for all sweeps, there is some variability on the maximal displacement across different parameter sweeps. The results are shown in Figure 9. Looking at the maximal displacement (top row), the order of magnitude is in line with [39] in which displacements of a couple of micrometers are measured. However, the simulated displacement in this study cannot be compared directly with [39], since different transducers and frequencies are used. Since the displacement field evolves over time, it is shown at two representative time points in Figure 9B-C: 10 ms, when the steady state is approximately reached, and 80 ms, corresponding to the end of the simulation. The metrics shown in Figure 9A are calculated after 80 ms.

**Fig. 9:**
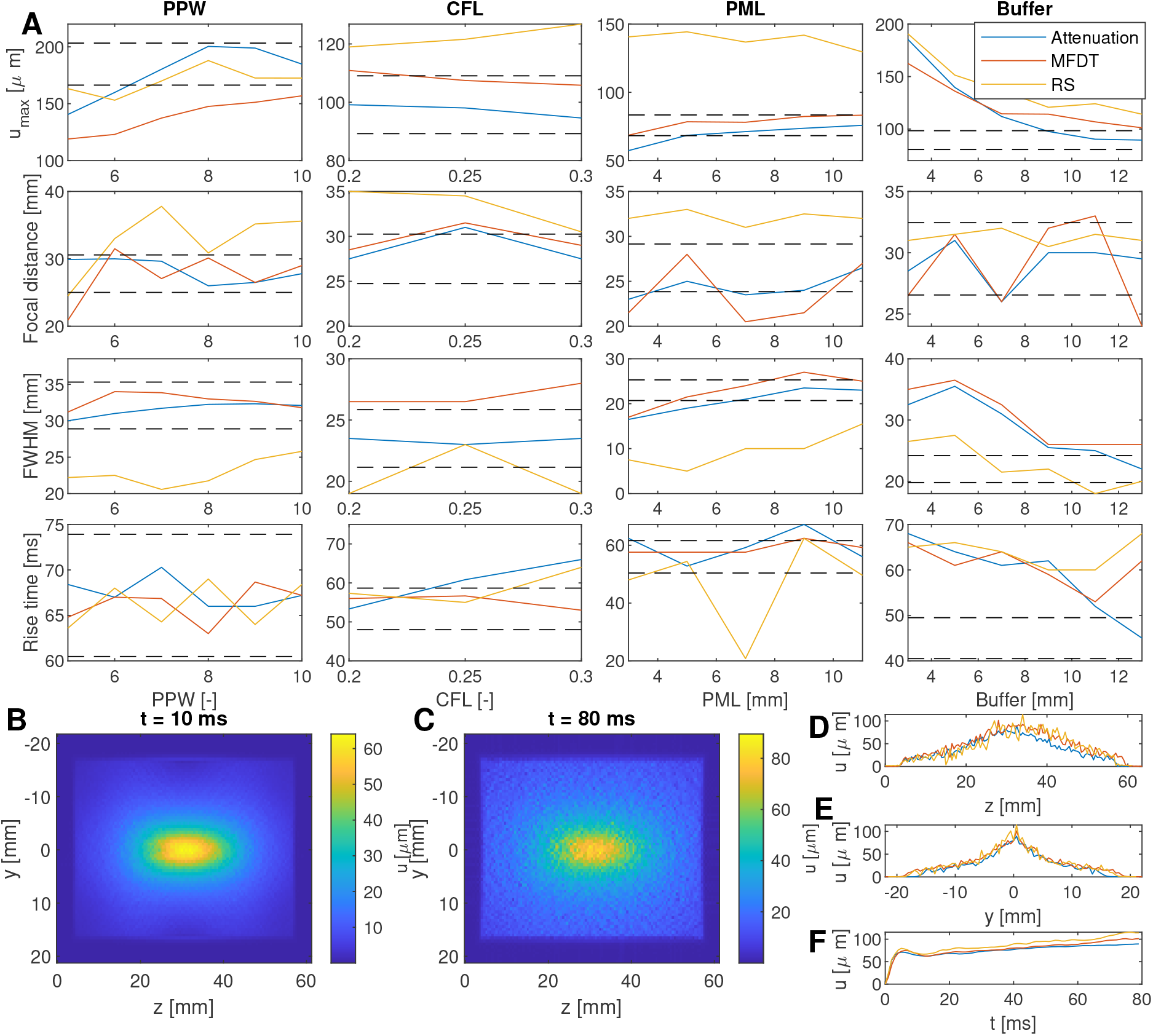
Illustration of the effect of simulation settings on displacement simulations in free space for a continuous ultrasound pulse of 80 ms. **A**: Influence of simulation parameters (PPW, CFL, PML, and buffer size) on metrics (maximal displacement magnitude (*u*_max_), focal distance, axial FWHM, and rise time) at *t* = 80 ms. 10 % error bars w.r.t. the most precise attenuation force simulation are indicated by black dashed lines. **B-C**: examples of 2D fields of displacement simulations with a continuous free space ARF input (attenuation formulation). **D**: Displacement on the axial axis after 80 ms. **E**: Lateral displacement after 80 ms. **F**: Time evolution of maximal displacement. **B-F** are the results for 6 ppw, a CFL of 0.3, a PML of 5 mm, and a buffer of 13 mm. All metrics are calculated on the axis through the maximum.

It can be seen in Figure 9A that most metrics do not follow a strictly increasing or decreasing trend (e.g., maximal displacement versus PPW, focal distance versus buffer, and rise time versus PPW). However, most metrics stay within the 10 % error margin for most of the sweep. For the maximal displacement as a function of the buffer zone, the metrics only fall within the 10 % error margin from 9 mm on for the attenuation formulation. For the FWHM and rise time of the displacement caused by the attenuation force as a function of the buffer zone, the metrics only fall within the 10 % error margin for the largest buffer zone. There is a clear downwards trend of the maximal displacement with increasing buffer size.

It can be observed from the plot at 80 ms (Fig. 9C), that the numerical noise in the displacement profiles increases with simulation time. This observation is related to reflections on the PML, that can be reduced by including the buffer region. The distance between the input force and the PML increases with buffer size. As a result, the displacement field reaching the PML decreases with the buffer size. Because k-Wave uses the Kelvin-Voigt model for the viscoelastic medium, it is expected that a steady state displacement will be reached. In Figure 9F, it can be seen that from 10 ms to 80 ms, a slow increase of 25 % is present. In the meantime, the noise is increasing as well, which seems to be inevitable across all tested simulation parameter settings. From this it can be concluded that the numerical requirements increase with simulation duration for ARF-driven displacement simulations. However, even for long sonication durations, the displacement simulations still provide a reasonable estimation of the displacement field for the application of ultrasound neuromodulation (Figure 9D-F).

When comparing the different ARF formulations as input for the displacement simulation, it can be seen that the resulting displacement distributions are close to each other (Figure 9D-F), even though the ARF is bigger for the RS formulation than for the attenuation and MFDT formulation (Figure 6). For the case with a buffer of 13 mm, the maximal displacement in Figure 9F caused by the attenuation force is 13 % and 28 % smaller than the displacement for the MFDT formulation and the RS formulation, respectively. For the hyperparameters used in Figure 9B-F and evaluated at 80 ms, the focal distance is 18 % higher than for the MFDT formulation and 5.1 % lower than for the RS formulation. The FWHM is 18 % lower than for the MFDT formulation and 9 % higher than for the RS formulation.

#### 2) Pulsed, skull

Here, the same analysis is repeated as in the previous section with the ARF of the realistic skull simulation of Figure 6 as input instead. The results are shown in Figure 10. It can be seen that PPW, CFL, and PML barely have an influence on the normalised maximal displacement, focal distance, FWHM, and rise time. Only the buffer layer goes above an error of 10 % for low values. However, there is a big difference between the ARF formulations for the maximal displacement. The attenuation formulation leads to a maximal displacement of 1.87 *µ*m, while the MFDT and RS formulations lead to a maximum displacement of 8.43 *µ*m and 8.39 *µ*m, respectively. Comparing the 2D and 1D plots (Figure 10B-F) to the continuous simulations (Figure 9B-F), it is clear that less noise is visible throughout the whole simulation for a pulsed insonication. As a result, the convergence with respect to different solver settings is improved for pulsed stimulation (Fig. 10A, 5 ms, realistic skull) compared to continuous insonication (Fig. 9A, 80 ms, free space).

**Fig. 10:**
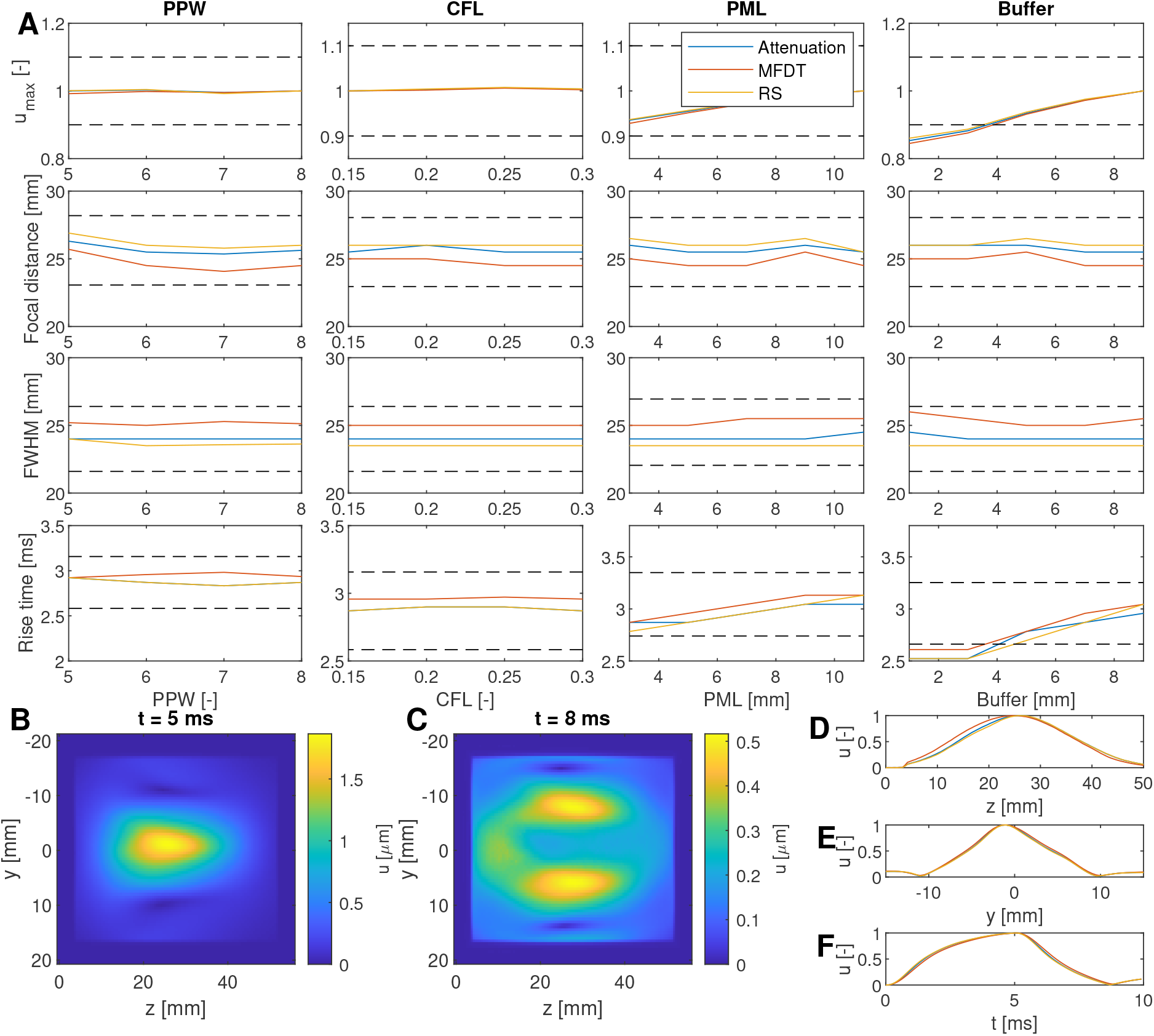
Illustration of the effect of simulation settings on displacement simulations with a realistic skull phantom for an ultrasound pulse of 5 ms, followed by an off-time of 5 ms. **A**: Influence of simulation parameters (PPW, CFL, PML, and buffer size) on metrics (maximal displacement (*u*_max_), focal distance, axial FWHM, and rise time) at *t* = 5 ms. The maximal displacement is normalised to the most accurate simulation setting for the respective ARF formulation for visualisation purposes. 10 % error bars w.r.t. the most precise attenuation force simulation are indicated by black dashed lines. **B-C**: examples of 2D fields of displacement simulations with a pulsed ARF input (attenuation formulation) for a pulse duration of 5 ms for the real skull ARF as input. **D**: Displacement on the axial axis after 5 ms, normalised to the maximum because the MFDT and RS lead to much higher displacement fields. **E**: Displacement on the lateral axis after 5 ms, normalised to the maximum. **F**: Time evolution of maximal displacement, normalised to the maximum. The maxima for the discretisation parameters in B-F are 1.87 *µ*m, 8.43 *µ*m, and 8.39 *µ*m for the attenuation formulation, MFDT formulation, and RS formulation, respectively. **B-F** are the results for 6 ppw, a CFL of 0.3, a PML of 5 mm, and a buffer of 9 mm. All metrics are calculated on the axis through the maximum.

## IV. Discussion

### A. ARF formulation and recommendations for numerical hyperparameters

From the results of the ARF formulations it is clear that for the free space and half skull cases, the attenuation force and MFDT formulation are close to each other. The RS formulation shows big differences with both other formulations. In the literature, there is a discussion of which formulation of the acoustic radiation force is preferred for the simulation of tissue displacement or acoustic streaming. It was stated in [19] that the attenuation formulation and MFDT formulation are equivalent for low attenuating media. Meanwhile, in [16] the choice for the RS formulation is motivated by its capacity to accurately model complex stress patterns. Finally, [18] argues that the MFDT is the most general formulation.

Our results indicate that the second order error *ϵ* of the attenuation force w.r.t. the MFDT formulation is negligible across all considered attenuation coefficients (Figure 7E) in accordance with the derivation of [19]. As a result, this study indicates that the attenuation force is preferred, considering the ease of computation and the reduction of numerical errors for coarses grids. Because it does not include spatial derivatives as the MFDT formulation does, a far less fine resolution of the grid is needed in order to calculate the ARF. Potentially, k-Wave is more susceptible to enlarging errors after taking the derivative because of the introduction of oscillations due to the use of Fourier transforms.

From this study it is clear that the attenuation force and MFDT converge to each other in free space for fine grid resolutions. However, it takes very fine resolutions for the MFDT formulations to really converge: 15 ppw for the MFDT formulation when taking the derivative via the gradient compared to 7 ppw for the attenuation formulation to have a numerical error on the maximal ARF below 5 %. The reason for this is that the two terms in the MFDT formulation ((14), Eulerian excess pressure and Reynolds stress) counteract each other and partially cancel each other out. Because of the derivatives in the equation, it takes fine grids for the Eulerian excess pressure and Reynolds stress term to properly cancel out each other to the right degree. Difference in simulation results for the heterogeneous cases show less clear results. The attenuation force is a lot smoother in any case, while the MFDT force follows a similar trend with more oscillations.

In summary, the following guidelines can be given. For pressure simulations that are used to subsequently calculate the ARF, ideally the settings mentioned in TABLE II can be used to minimize the numerical error. The PML can be reduced to 2 mm in a homogeneous simulation and 4 mm in a heterogeneous simulation. The grid resolution can be reduced to 7 ppw when using the attenuation formulation or MFDT using the k-space derivative in a homogeneous medium. In case of a quick simulation or a situation where computational resources are limited, less precise settings can be used to obtain a first impression of the field distribution, although this will result in higher errors. In particular, a lower number of ppw can reduce the computational requirements. In this case, it is still advised to use at least 7 ppw for the heterogeneous case.

The attenuation force is the most reliable and gives accurate results for all simulation scenarios (e.g., medium and transducer geometry). The error between the MFDT and attenuation force increases with transducer curvature, while it is less sensitive to the aperture, in agreement with the larger pressure field spatial gradients at a lower radius of curvature.

The displacement simulations should be used for short simulation pulses: from the moment a wave has time to travel across the whole simulation domain, interference with reflections on the boundaries of the simulation domain occurs and the error starts to increase: 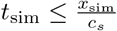, where *t*_sim_ is the maximal simulation time, *x*_sim_ is the size of the simulation domain and *c*_*s*_ is the shear wave speed. This can be mitigated by adding a buffer region to the simulation domain with the same medium properties in between the ARF sources and the PML.

Because of the linearity of the solver, the pressure fields can be scaled linearly with the transducer pressure. The ARF and displacement fields should be scaled quadratically with the transducer pressure.

### B. Limitations and future work

For the calculation of the acoustic radiation force, a quasistatic steady-state approach is used (i.e., transient effects are neglected). In reality, a pulsed protocol is used and the ARF has a specific rise time. In order to correctly model this, the time step of the simulation will have to be analysed more in depth.

This study focused specifically on the optimization of the accuracy of simulations in k-Wave because the k-Wave toolbox is widely used in computational research for ultrasonic neuromodulation, as well as for treatment planning in the form of k-Plan [17], [18], [40], [41], [42]. In future work, it would be interesting to continue this research by investigating other solvers. k-Wave is established as a fast pseudospectral solver and its accuracy is confirmed in [15] when it comes to the simulation of the pressure field. It would be valuable to expand this validation to ARF and displacement fields as well, e.g., with shear wave elastography, ultrasound displacement imaging or magnetic resonance acoustic radiation force imaging (MR-ARFI) [43], [8], [44]. For the calculations of the ARF where the derivative is taken in the k-space, it can be seen (Figure 5C) that the MFDT formulation follows the same trend as the attenuation formulation, however, with strong oscillations. Possibly, with accurate filtering techniques to reduce the oscillatory noise, the MFDT could still be used with a more coarse grid [45]. The displacement simulations are computationally heavy because of the stress tensors included in the calculation. For this case specifically, the simulation time was the limiting factor. This reduces the amount of freedom to investigate all parameters, and as a result the default parameters are not fixed across parameter sweeps in Figs. 9-10 (cf. TABLE III). For example, it is not possible to include the whole simulation domain and to calculate the evolution of the displacement for several milliseconds. However, a longer simulation time is necessary because of the slow evolution of the displacement with respect to the pressure, that rises within a couple of microseconds. Moreover, when including skull tissue in the displacement simulations, the time step needs to be at least six times smaller which reduces the freedom in parameter choice even further. Additionally, it was seen that for long pulses, the numerical noise in the displacement profiles will increase over the simulation duration. Because of this, displacement simulations should be restricted to short pulses. In this case, a pulse of 5 ms was chosen. Looking at typical TUS protocols, this can be considered as a relatively long pulse already [46]. Only a handful of studies are found in which considerably longer pulses are used [47], [48].

## V. Conclusions

In this study, numerical methods for calculating ARF and tissue displacement are discussed. It is concluded that the attenuation force formulation is optimal for calculating the ARF. The main reason is that it is numerically easier to compute: it does not include derivatives that magnify the noise from the pressure simulation, specifically from the Gibbs phenomenon due to Fourier transforms in k-Wave. Moreover, in the MFDT formulation, parts of both terms (Eulerian excess pressure and Reynolds stress) only cancel out at high simulation grid resolutions due to the slow convergence of the derivatives. In some cases, as in homogeneous simulation domains, one can still opt to use the MFDT formulation. In this case it is recommended to calculate the spatial derivatives in the k-space. This is the same method that is used to compute spatial derivatives in the k-Wave simulation itself. Futhermore, it was found that the tissue attenuation has a limited effect on the choice of the ARF calculation method. Only for low attenuation coefficients, there is a discrepancy. In this case, a higher grid resolution is needed to get accurate results. This is a result of increasing numerical errors with decreasing attenuation, and is not due to the physics of this problem: in the whole tested range of attenuation coefficients, the attenuation formulation should be equivalent to the MFDT formulation, as long as the grid resolution is fine enough.

In free space, all ARF formulations result in approximately the same displacement distribution with a sharp increase at the onset of the ultrasound pulse, reaching a plateau after a couple of milliseconds. When adding a realistic skull, the displacement distributions differ more because of the differences in ARF distribution. Moreover, it was observed that noise builds up in the displacement simulation result over time. This can be mitigated by adding a buffer layer, keeping the ARF sources further away from the simulation boundary. Because of the build up of noise during the displacement simulation, it is recommended to restrict the simulation duration to the shear wave transit time through the simulation domain.

## Data Availability

All data produced are available online at 10.5281/zenodo.23976809

## VI. Data availability

The code used in this paper is available on https://github.com/toplovie/UNMODARFFormulation.

The Data used for the plots is available on 10.5281/zenodo.23976809

## Notes

### Competing Interest Statement

The authors have declared no competing interest.

### Author Declarations

This work used simulated data and data from https://www.kaggle.com/datasets/fooxmoon/cq500-dataset

### Funding

- Fonds Wetenschappelijk Onderzoek (FWO): G019121N, G0A5623N

