## Supplementary text for "Computational Modeling of the Acoustic Radiation Force and Viscoelastic Displacement for Transcranial Focused Ultrasound Neuromodulation"

#### I. Equivalence of MFDT and attenuation force formulations at low attenuation coefficients

The MFDT and attenuation force formulations are identical for low attenuation coefficients. An upper bound on the attenuation coefficient can be derived as a condition of validity for the equivalence between both ARF formulations, starting from the derivation in [1]. The linearized Navier-Stokes equation for irrotational flow is

$$\rho_0 \frac{\partial \boldsymbol{v}}{\partial t} = -\nabla p + \left(\zeta + \frac{4}{3}\eta\right)\nabla(\nabla \cdot \boldsymbol{v}), \tag{1}$$

where $\rho_0$ is the density, $\boldsymbol{v}$ is the velocity, $p$ is the thermodynamical pressure, $\zeta = \chi + \frac{2\eta}{3}$ is the bulk viscosity, and $\eta$ is the shear viscosity. This generalizes the k-Wave equations ((1)-(2) in the paper) by including viscosity terms. The continuity equation and equation of state are:

$$\frac{\partial \rho}{\partial t} + \rho_0 \nabla \cdot \boldsymbol{v} = 0, \tag{2}$$

$$p = c_0^2 \rho, \tag{3}$$

Using phasor notation, the following expression for the complex amplitudes is retrieved by combining (1), (2), and (3):

$$V_i = \left(\frac{1}{\rho_0 j\omega} - \frac{\zeta + \frac{4}{3}\eta}{\rho_0^2 c_0^2}\right)\frac{\partial P}{\partial x_i}. \tag{4}$$

This equation can also be written as:

$$\nabla P = \gamma(\alpha_p)\boldsymbol{V} \tag{5}$$

Here, $\alpha_p(\omega) = \frac{(\zeta + 4/3\eta)\omega^2}{2\rho_0 c_0^3}$ is the attenuation coefficient and $\gamma(\alpha_p)$ is given by:

$$\gamma(\alpha_p) = \frac{\rho_0 j\omega^2}{\omega - 2j\alpha_p(\omega)c_0} = \rho_0 j\omega - 2\rho_0 c_0 \alpha_p(\omega) + \mathcal{O}(\alpha_p(\omega)^2) \tag{6}$$

The MFDT ARF formulation can be written as [1]:

$$\boldsymbol{F}_{\text{MFDT}} = \frac{j\alpha_p(\omega)}{2c_0\rho_0\omega}(P\nabla P^* - P^*\nabla P) \tag{7}$$

Substituting (5) back in (7), the ARF can also be written as:

$$\boldsymbol{F}_{\text{MFDT}} = \overbrace{\frac{\alpha_p(\omega)}{2c_0}(\boldsymbol{V}^*P + \boldsymbol{V}P^*)}^{\boldsymbol{F}_1} + \underbrace{\frac{\alpha_p(\omega)^2}{\rho_0\omega^2}(P\nabla P^* + P^*\nabla P)}_{\boldsymbol{F}_2} + \mathcal{O}(\alpha_p^3(\omega)). \tag{8}$$

The first order term is then equal to

$$\boldsymbol{F}_1 = \frac{2\alpha_p(\omega)\langle \boldsymbol{I}\rangle}{c_0}. \tag{9}$$

The second order ARF is given by

$$\boldsymbol{F}_2 = \frac{\alpha_p(\omega)^2}{\rho_0\omega^2}(P\nabla P^* + P^*\nabla P). \tag{10}$$

As a result, the second order term is small enough when (9) is much bigger than (10). This is equivalent to:

$$\alpha_p(\omega) \ll \frac{2I\rho_0\omega^2}{c_0||P\nabla P^* + P^*\nabla P||}. \tag{11}$$

#### II. The MFDT and RS formulations induce identical acoustic streaming and viscoelastic displacement distributions

The relationship between the ARF in the MFDT and RS formulation is:

$$\boldsymbol{F}_{\text{MFDT}} = -\nabla p_{\text{E}} + \boldsymbol{F}_{\text{RS}}, \tag{12}$$

where $\boldsymbol{F}_{\text{MFDT}}$ is the MFDT formulation, $p_{\text{E}}$ is the Eulerian excess pressure, and $\boldsymbol{F}_{\text{RS}}$ is the RS formulation. When using the RS formulation as an input to calculate acoustic streaming, the Navier-Stokes equation becomes in second order [2]:

$$\rho_0\frac{\partial \boldsymbol{v}^{(2)}}{\partial t} = -\nabla p_2 + \left(\zeta + \frac{4}{3}\eta\right)\nabla(\nabla\cdot\boldsymbol{v}^{(2)}) - \eta\nabla\times\nabla\times\boldsymbol{v}^{(2)} + \boldsymbol{F}_{\text{RS}}, \tag{13}$$

where $\boldsymbol{v}^{(2)}$ and $p_2$ are the second order velocity and pressure in the second simulation step, respectively. Combining this with (12), we obtain:

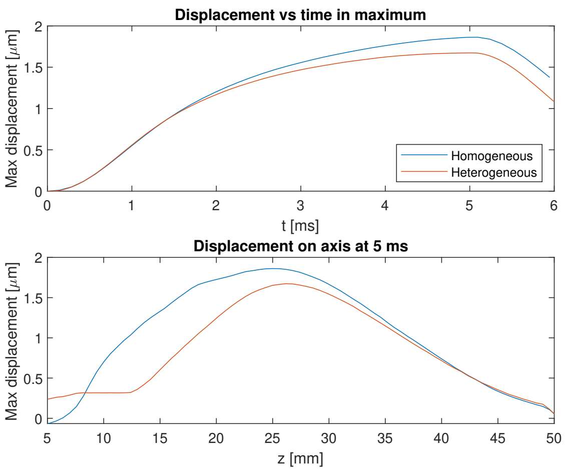


Fig. 1. Effect of including the skull in the displacement simulation, with the transducer located at $z = 0$ mm. A shift of the maximal displacement away from the skull and a decrease in FWHM can be observed. The simulations are done for a grid resolution of 5 ppw, a PML thickness of 5 mm, and a buffer of 5 mm. The CFL was 0.3 for the homogeneous simulation and 0.05 for the heterogeneous simulation. The input of the displacement simulation is the attenuation ARF for the realistic skull (5 ms pulse).

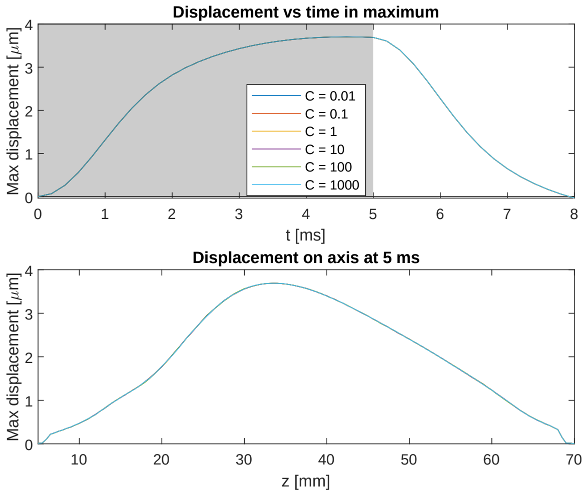


Fig. 2. Effect of the shear wave attenuation on the displacement for the homogeneous case, showing it has minimal effect. Top figure shows the maximal displacement over time. The grey area indicates the stimulation time. Bottom figure shows the displacement on the main axis after 5 ms. C is the ratio of shear wave attenuation and compressional wave attenuation. The simulations are done for a grid resolution of 6 ppw, a CFL of 0.3, PML thickness of 5 mm, and a buffer of 5 mm.

$$\rho_0 \frac{\partial \boldsymbol{v}^{(2)}}{\partial t} = -\nabla(p_2 - p_\mathrm{E}) + \left(\zeta + \frac{4}{3}\eta\right)\nabla(\nabla\cdot\boldsymbol{v}^{(2)}) - \eta\nabla\times\nabla\times\boldsymbol{v}^{(2)} + \boldsymbol{F}_\mathrm{MFDT}. \quad (14)$$

This equation is solved, together with the continuity equation, which is in second order:

$$\rho_0\nabla\cdot\boldsymbol{v}^{(2)} + \nabla\cdot\langle\rho\boldsymbol{v}\rangle = 0 \quad (15)$$

From this, it can be observed that only $p_2$ changes when using a different formulation, while the acoustic streaming velocity $\boldsymbol{v}^{(2)}$ is identical between both ARF formulations.

Under the assumption of incompressibility, a similar reasoning can be followed for a Kelvin-Voigt material, used in this study [3]:

$$\sigma_{ij} = \lambda\delta_{ij}\epsilon_{kk} + 2\mu\epsilon_{ij} + \chi\delta_{ij}\frac{\partial\epsilon_{kk}}{\partial t} + 2\eta\frac{\partial\epsilon_{ij}}{\partial t} \quad (16)$$

$$= \underbrace{\left(\kappa\epsilon_{kk} + \zeta\frac{\partial\epsilon_{kk}}{\partial t}\right)}_{-p_2}\delta_{ij} + 2\mu\hat{\epsilon}_{ij} + 2\eta\frac{\partial\hat{\epsilon}_{ij}}{\partial t} \quad (17)$$

With $\kappa = \lambda + \frac{2\mu}{3}$ the bulk modulus. $\epsilon_{ij} = \frac{1}{2}(\frac{\partial u_i}{\partial x_j} + \frac{\partial u_j}{\partial x_i})$ is the strain tensor, $\hat{\epsilon}_{ij} = \epsilon_{ij} - \frac{1}{3}\delta_{ij}\epsilon_{kk}$ is the deviatoric strain tensor, and $\boldsymbol{u}$ is the displacement ($\frac{\partial\boldsymbol{u}}{\partial t} = \boldsymbol{v}$). As for the fluid, the conservation of momentum equation and the incompressibility condition ($\epsilon_{kk} = \nabla\cdot\boldsymbol{u} = 0$), are solved together with the constitutive equation (17). The excess pressure $p_\mathrm{E}$ can then again be absorbed in the second order pressure $p_2$.

### III. Displacement simulations

#### A. Including skull tissue influences the tissue displacement at a high computational cost

Figure 1 shows the effect of including the skull in the displacement simulation. It can be seen that the focal spot shifts away from the skull. Note that only the CFL is changed between both simulations, in order to keep the simulation stable. Because these simulations are computationally intensive, the grid resolution is kept constant at 5 ppw, which is a relatively low spatial discretisation resolution and is expected to impact the numerical accuracy.

#### B. Shear wave attenuation of the brain does not influence the resulting displacement

The shear wave speed and attenuation in the skull are set to $(1400/2550)c_{p,\mathrm{skull}}$ and $(90/85)\alpha_{p,\mathrm{skull}}$, respectively, as reported in [4]. In [4], however, the shear waves are used in the pressure simulation, while in this study, the shear waves are only included in the second order simulation to calculate the tissue displacement. Estimations of the shear wave attenuation in the brain can be made from the loss modulus [5]. However, to our knowledge, no literature directly reports shear wave attenuation in the brain. For this reason, it is checked how much the shear wave attenuation influences the simulation result. The shear wave attenuation is varied in relation to the compressional wave attenuation coefficient ($\alpha_{s,\mathrm{brain}} = C\alpha_{p,\mathrm{brain}}$). In Figure 2, the shear wave attenuation is changed from $C = 0.01$ to $C = 1000$. However, the shear wave attenuation in the brain barely impacts the result, which is attributed to the difference in order of magnitude between the shear and compressional wave speed. In the paper, the shear wave attenuation is taken equal to the compressional wave attenuation.
